## Supplementary material for "Assessing the spatial-temporal risks of SARS-CoV-2 infection for healthcare-workers in the hospital using behavioural indices from routine data"

### *Methods*

#### *Validation of shift identification*

At the time of data extraction the rostering data for different staff groups were not on a centralised system. The roster data that was accessible did not include key staff roles such as doctors, and primarily covered nurses and healthcare assistants. We therefore inferred shifts using the routinely collected data to ensure shift metrics and key occupational roles of interest could be included in the study. We used the available rostering data (during the observation period) to validate our methods for inferring shift metrics; to decide the most appropriate lag time between events for identifying a new shift and methods for identifying night shifts.

The rostering data was first cleaned by removing any shifts that were not in the UCLH Tower building, and shifts that were considered extensions of an already existing shift on the same day e.g. shifts entered as over time. In total, the cleaned rostering data contained information on 394,611 shifts. Of these, 233,242 were labelled as working (i.e. not days off or annual leave), of which 78% had evidence of activity (door events and/or patient contacts logged in the Tower building by the individual) during the shift. Shifts without evidence for HCW activity will be due to either HCWs not logging any events in the Tower building during the shift or errors in the rostering data set e.g. 4% of shifts labelled as not working had evidence of activity in the events data.

In the available rostering data the earliest start time of a day shift was 6am, while the earliest start time for a night shift was 18:00. For this reason we identified night shifts using the time of the first event, whereby an event between 17:00 and 5am identified a night shift. We used an hour buffer either side of the observed start times as it was reasonable to assume the first logged event of a shift may be earlier than the start time if individuals turned up early.

To determine the most appropriate lag time between events for identifying new shifts, we compared the results of using either a 4 hour, 7.5 hour or 11 hour lag. We calculated the sensitivity and degree of errors produced by each method. Sensitivity was calculated as the percentage of working shifts in the rostering data (with evidence of activity) that shared a start date with shifts identified using the routinely collected data. We then calculated the percentage of working rostered shifts (with evidence of activity) that were associated with errors in shifts derived from the routinely collected data. These errors included either (1) multiple shifts identified from the routinely collected data that shared the same start date (indicating an over estimation in the number of shifts) or (2) errors in determining the end date of a shift i.e. day shifts with more than one date associated with them or night shifts with more than two dates. As seen in Table S1, a lag of 7.5 hours was the most effective method of correctly identifying rostered shifts with the fewest errors.

**Table S1. Lag time between events used to identify new shifts.** A lag time of either 7.5 hours, 4 hours or 11 hours was used to identify rostered shifts in routinely collected data (patient contacts and door events) at UCLH. For working rostered shifts with evidence of activity, the percentage of all shifts and night shifts identified in the events data is reported. For rostered shifts detected in the events data, the percentage is reported for those with errors due to (1) multiple shifts being identified on the same day and (2) incorrect end dates being identified (day shifts with >1 date associated with them or night shifts with >2 dates).

| | 7.5 hrs | 4 hrs | 11 hrs |
|---|---|---|---|
| All working rostered shifts identified in the events data | 99% | 100% | 94% |
| Working rostered night shifts identified in the events data | 97% | 90% | 99% |
| Errors due to multiple shifts with the same start date in the events data | 3% | 18% | 1% |
| Errors due to day shifts in the events data having an incorrect end date | <1% | <1% | 4% |
| Errors due to night shifts in the events data having an incorrect end date | <1% | <1% | 7% |

*Directed acyclic graph*

In causal inference, the aim is to estimate the true causal association between an exposure variable and an outcome variable. This is done by removing (usually via statistical adjustment) all other hypothesized associations that confound the focal relationship.

Directed acyclic graphs (DAGs) are graphical models that map out the relationships between explanatory variables and the outcome variable. A DAG can be interpreted as a graphical representation of hypotheses about the data-generating process and provide a summary of the modelling approach. For an introduction to the formal causal inference framework and DAGs see [1–3].

For our analyses, we used logistic regression with mixed effects to investigate the total causal effect of various exposures on the likelihood of healthcare workers (HCWs) testing positive for COVID-19. Exposures of interest include ethnicity, age, staff role, shift patterns, patient contacts and HCW mobility (Figure S1).We extracted information from routinely-collected hospital data sources for each individual who completed a COVID-19 test.

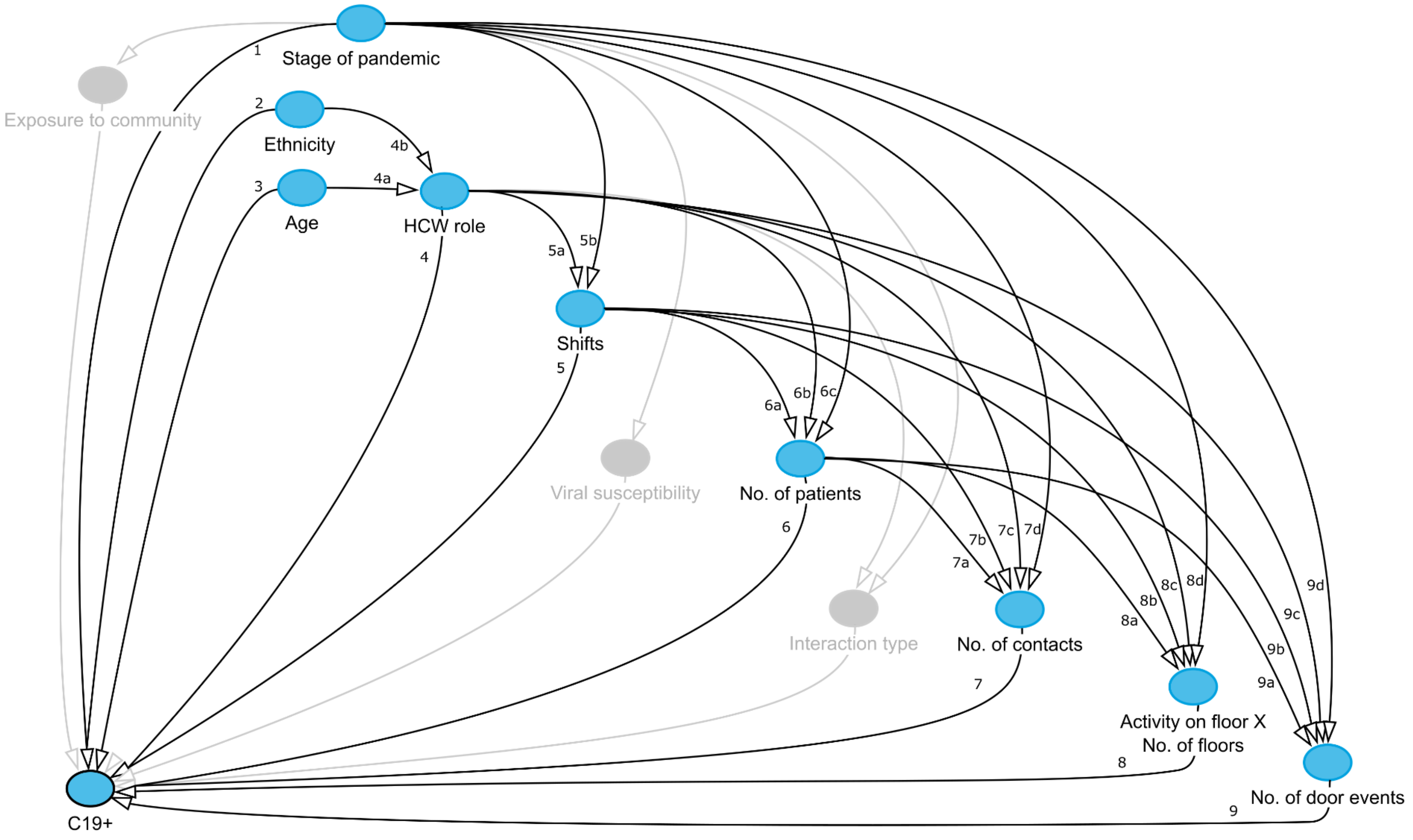


*Figure S1. Directed acyclic graph (DAG). The DAG depicts the hypothesised relationships between variables considered to influence the probability of healthcare workers testing positive for COVID-19 (C19+). The blue nodes represent the observed variables while the grey nodes represent unobserved variables. Arrows connecting nodes show the (directional) relationships between variables. The numbers next to lines refer to the notations section; that provides justifications for the hypothesised relationships.*

*Figure S1 notations; Hypothesised associations between variables*

1. **Stage of the pandemic**
   - Hypothesis: The probability of HCWs testing positive for COVID-19 will change as the pandemic progresses, and will be highest during the first wave.
   - Supporting evidence/theory: Based on the changing number of COVID-19 patients in hospital, the pandemic can be split into distinct stages (see methods of main article). The probability of HCWs testing positive for COVID-19, will not be equal across the different stages of the pandemic, owing to multiple unobserved factors including differences in hospital admissions, community transmission, PPE availability and hospital and community interventions.

2. **Ethnicity**
   - Hypothesis: HCWs in the BAME ethnic group will by more likely to have a positive test result than other ethnicities.
   - Supporting evidence/theory: As discussed in the review by Out et al. (2020)[4], in the UK and during the early stages of the pandemic, there was growing evidence that Black, Asian and Minority Ethnicities (BAME) were disproportionately affected by COVID-19 than white ethnicities. The BAME ethnic group were more likely to be diagnosed with COVID-19 and to exhibit severe disease. These health inequalities are thought to arise from a complex network of socio-economic factors; income, social class, occupational background and educational achievement.

3. **Age**
   - Hypothesis: The probability of a HCW testing positive for COVID-19 will increase with HCW age.
   - Supporting evidence/theory: There is evidence to suggest that the susceptibility to infection increases with age [5,6].

4. **Healthcare worker (HCW) role**
   - Hypothesis: Doctors, nurses and healthcare assistants will have higher probabilities of testing positive for COVID-19 than other staff groups.
   - Supporting evidence/theory: Doctors, nurses and healthcare assistants have been consistently identified as occupations with a higher risk of testing positive for COVID-19 [7–14].
   - Adjusted variables:
     a. Age; The role of HCWs may depend on their age, particularly those roles that require a high level of experience and training.
     b. Ethnicity; In the UK, an individuals ethnicity can influence their occupation and, in the NHS, the BAME ethnic group are less likely to occupy more senior HCW roles [15].

5. **Shifts**
   - Hypothesis: The probability of a HCW testing positive for COVID-19 will increase with (i) the number of shifts they work and (ii) the relative number of night shifts they work.
   - Supporting evidence/theory: In a study by Maidstone et al. (2021), that used data from the UK Biobank, individuals considered to be an essential worker and those with shift work were more likely to test positive for COVID-19. Another study using data from the

UK Biobank found that night shifts were associated with higher risks of testing positive for COVID-19 [17]. Similar results were found for HCWs working at a hospital in Italy [18].

- Adjusted variables:
    - **a.** HCW role; The rostering of staff may be determined by their occupational role. Ethnicity and age are assumed to exert no direct influence on shift patterns beyond the impact they have on determining HCW role.
    - **b.** Stage of the pandemic; Different stages of the pandemic may require individuals to work more/less shifts due to staff shortages, IPC interventions or increased hospital pressures.

**6. Patient engagement; Number of patients**

- Hypothesis: The probability of a HCW testing positive for COVID-19 will increase with (i) the total number of patients they interact with and (ii) the relative number of COVID-19 patients they interact with.
- Supporting evidence/theory: One of the main pathways for the transmission of communicable diseases such as SARS-CoV-2, is through direct contact (within 2m) with an infectious individual [19]. HCWs can be exposed to both patients known to have COVID-19 and those who are not known to have the virus but are asymptomatic or pre-symptomatic [20]. There is evidence in the literature for an increased risk of HCWs testing positive if they are patient facing or in contact with COVID-19 patients [10,11,21]. The number of patients and/or number of COVID-19 patients that a HCW interacts with may therefore provide a measure of exposure to the virus in the hospital.
- Adjusted variables:
    - **a.** Number of shifts; The number of patients a HCW sees will be positively associated with the number of shifts they work.
    - **b.** HCW role; The number of patients seen by HCWs will depend on their occupation, with some roles interacting with more patients than others [22,23]. Ethnicity and age are assumed to exert no direct influence on patient engagement beyond the impact they have on determining HCW role.
    - **c.** Stage of the pandemic; The number of patients seen by a HCW will depend on the number of patients in the hospital, and this will be influenced by the stage of the pandemic.

**7. Patient engagement; Number of patient contacts**

- Hypothesis: The probability of a HCW testing positive for COVID-19 will increase with (i) the total number of patient contacts they have and (ii) the relative number of contacts they have with COVID-19 patients.
- Supporting evidence/theory: For communicable diseases, the frequency of contact between infectious and susceptible individuals is known to be an important determinant for the risk of transmitting and acquiring infection [24,25]. There is some evidence that HCWs who have more frequent contact with COVID-19 patients in hospital are more likely to test positive for the disease [7,9].
- Adjusted variables:
    - **a.** Number of patients; The number of patient contacts a HCW has will be positively associated with the number of patients they interact with.
    - **b.** Number of shifts; The number of shifts a HCW works will be positively associated with the number of patient contacts they have.

c. HCW role; The number of patient contacts will depend on the HCWs occupation, with roles varying in their level of patient engagement [26,27]. Ethnicity and age are assumed to exert no direct influence on patient engagement beyond the impact they have on determining HCW role.

d. Stage of the pandemic; The number of patient contacts will depend on the stage of the pandemic due to changes in the patient population and associated needs for care. This was the case on a geriatric unit in France during an influenza epidemic, whereby the number of contacts between patients and nurses were higher outside of the epidemic [28].

**8. Mobility; Activity on floor X & Number of floors**

- Hypothesis: The probability of a HCW testing positive for COVID-19 will (i) not be equal across floors, (ii) depend on the number of floors they are active on and (iii) the relative number of COVID-19 floors they are active on.
- Supporting evidence/theory: The spatial movements of individuals can determine their exposure to pathogens. During the pandemic HCWs were reported to be more likely to test positive for COVID-19 if they were active on non-emergency wards [14] and on COVID-19 wards [9,12,13]. The number of floors an individual is active on provides a proxy for mobility in terms of the diversity of locations visited, while the number of COVID-19 floors provides an indication of an individuals mobility between COVID-19 hotspots. We assume that a HCWs movements around the hospital are determined by their occupation and the patients they are required to see (as opposed to their spatial movements determining the patients they see).
- Adjusted variables:

  a. Number of patients; The number of patients an individual sees will influence the spatial activity of HCWS, whereby the spatial distribution of patients will scale with the number of patients staff need to see (resulting from the constrained hospital layout). We hypothesised no direct relationship between an individuals number of patient contacts and their activity on particular floors or the number of floors they were active on.

  b. Shifts; The number of shifts an individual works will influence the number of floors they are active on, as there will be a greater likelihood of being rostered to multiple wards.

  c. HCW role; A HCWs occupation may determine the wards/departments they work on, and therefore the floors they are active on. Ethnicity and age are assumed to exert no direct influence on within hospital mobility beyond the impact they have on determining HCW role.

  d. Stage of the pandemic; The stage of the pandemic will determine the floors HCWs are active on, because the function of wards and their accessibility (closed/open) will depend on the changing circumstances of the pandemic.

**9. Mobility; Number of door events**

- Hypothesis: The probability of a HCW testing positive for COVID-19 will be influenced by (i) the number of door events they have and (ii) the relative number of door events on COVID-19 floors.
- Supporting evidence/theory: The number of door events provides a proxy for mobility in terms of the frequency of movement between spatially distinct areas (floors/wards). More movement events may be associated with more risky behaviours, such as the need

to don and doff PPE. Equally, a higher frequency of movement events may imply transient activities that reduce an individuals exposure to the virus.

- Adjusted variables:
    **a.** Number of patients; The number of patients an individual sees will influence the number of floors they are active on, and therefore their number of door events. We hypothesised no direct relationship between an individuals number of door events and number of patient contacts.
    **b.** Shifts; The number of door events will scale with the number of shifts worked.
    **c.** HCW role; A HCWs occupation may determine the wards/departments they work on, and their mobility around the hospital. Ethnicity and age are assumed to exert no direct influence on within hospital mobility beyond the impact they have on determining HCW role.
    **d.** Stage of the pandemic; The stage of the pandemic will determine the mobility of HCWs, as the function of wards and their accessibility (closed/open) will depend on the changing circumstances of the pandemic.

*A note on unobserved confounding variables*

Inferring causal effects can be difficult when handling data from non-randomised studies and when using retrospective observational data [29,30]. This arises from various sources of bias referred to as confounding e.g. selection bias, concurrent events and lack of comparability across groups.

We have attempted to control for the confounding effects of each exposure variable on the outcome, however, there is likely to be confounding due to unobserved variables (exposure to the virus in the community, viral susceptibility and hospital interventions) which we have either identified in our DAG (in grey) or tried to control for using the temporal covariate ‘stage of the pandemic’. Moreover, our dataset may contain unobserved sampling biases and, to reduce the complexity of the DAG, we have not included these variables in the figure but instead explicitly state them below:

(1) Testing in time. The hospital’s staff testing programme was rolled out when the pandemic was announced. In the first wave this was for symptomatic staff and after May 2020 it included weekly testing of asymptomatic staff. The covariate ‘stage of the pandemic’ should control for the different testing policies, however we are unable to control for other related sources of bias such as compliance (given tests were taken at the HCWs own discretion) or tests taken outside the staff testing programme.

(2) Routinely collected data. We excluded test results for individuals that had no recorded patient contacts and door events in the routinely collected hospital data. Sampling bias may be generated through variation in the way HCWs record patient contacts or use security doors e.g. during ward rounds consultants move from patient to patient with groups of JR doctors following (risk of tail-gating), one of which will write up notes; this could result in biases between staff roles in the calculation of behavioural metrics

Table S2. Model terms. All models were mixed effects logistic regressions, and included individual ID as a random effect.

| Variable of interest | Formula |
|---|---|
| Stage of pandemic / Ethnicity / Age | Stage + Ethnicity + Age |
| HCW role | Role*Stage + Ethnicity + Age |
| *Shifts* | |
| No. of shifts | Shifts*Stage + Role*Stage |
| No. of night shifts (relative) | Nights*Stage + Shift*Stage + Role*Stage |
| *Patient contacts* | |
| No. of patients | Patients*Stage + Shifts*Stage + Role*Stage |
| No. of COVID-19 patients (relative) | PatientsC19*Stage + Patients*Stage + Shifts*Stage + Role*Stage |
| No. of patient contacts | Contacts*Stage + Patients*Stage + Patients*Stage + Shifts*Stage + Role*Stage |
| No. of COVID-19 contacts (relative) | ContactsC19*Stage + Contacts*Stage + PatientsC19*Stage + Shifts*Stage + Role*Stage |
| *Mobility* | |
| Activity on floor X (Y/N) | FloorX*Stage + Patients*Stage + Shifts*Stage + Role*Stage |
| No. of floors | Floors*Stage + Patients*Stage + Shifts*Stage + Role*Stage |
| No. of COVID-19 floors (relative) | FloorsC19*Stage + Floors*Stage + PatientsC19*Stage + Shifts*Stage + Role*Stage |
| No. of door events | DoorEvents*Stage + Floors*Stage + Patients*Stage + Shifts*Stage + Role*Stage |
| No. of door events on COVID-19 floors (relative) | DoorEventsC19*Stage + DoorEvents*Stage + FloorsC19*Stage + PatientsC19*Stage + Shifts*Stage + Role*Stage |

Table S3: Post-hoc comparisons for the risk of testing positive for COVID-19 between healthcare worker roles during the first wave. The probability of testing positive for COVID-19 is reported for each healthcare worker role in the diagonal of the matrix (grey cells) with 95% confidence intervals in brackets. The contrasts between roles, as determined by estimated marginal means from a mixed effects logistic regression, are reported as odds ratios with 95% confidence intervals in brackets. Contrasts with statistical significance are in bold.

| | First wave | Admin | Allied health professional | Doctor: consultant | Doctor: trainee | Healthcare assistant | Nurse | Other: clinical | Physiotherapist |
|---|---|---|---|---|---|---|---|---|---|
| *Reference level* | Admin | 14% (7-26) | | | | | | | |
| | Allied health professional | 1.08 (0.27-4.31) | 13% (8-20) | | | | | | |
| | Doctor: consultant | 1.90 (0.43-8.31) | 1.76 (0.56-5.49) | 8% (4-14) | | | | | |
| | Doctor: trainee | 0.92 (0.23-3.68) | 0.85 (0.32-2.30) | 0.49 (0.15-1.53) | 15% (10-23) | | | | |
| | Healthcare assistant | 1.10 (0.28-4.28) | 1.01 (0.38-2.72) | 0.58 (0.19-1.77) | 1.19 (0.44-3.19) | 13% (8-20) | | | |
| | Nurse | 1.33 (0.39-4.51) | 1.23 (0.57-2.64) | 0.7 (0.27-1.8) | 1.44 (0.67-3.09) | 1.21 (0.58-2.56) | 11% (8-14) | | |
| | Other: clinical | 1.43 (0.34-6.06) | 1.32 (0.44-3.96) | 0.75 (0.22-2.55) | 1.54 (0.51-4.64) | 1.30 (0.44-3.82) | 1.07 (0.44-2.61) | 10% (6-17) | |
| | Physiotherapist | 0.60 (0.10-3.70) | 0.55 (0.12-2.58) | 0.31 (0.06-1.63) | 0.64 (0.14-3.02) | 0.54 (0.12-2.55) | 0.45 (0.11-1.83) | 0.42 (0.08-2.12) | 22% (10-40) |

*p < 0.05; **p < 0.01; ***p < 0.001

Table S4: Post-hoc comparisons for the risk of testing positive for COVID-19 between healthcare worker roles during the summer lull. The probability of testing positive for COVID-19 is reported for each healthcare worker role in the diagonal of the matrix (grey cells) with 95% confidence intervals in brackets. The contrasts between roles, as determined by estimated marginal means from a mixed effects logistic regression, are reported as odds ratios with 95% confidence intervals in brackets. Contrasts with statistical significance are in bold.

*Reference level*

| Summer lull | Admin | Allied health professional | Doctor: consultant | Doctor: trainee | Healthcare assistant | Nurse | Other: clinical | Physiotherapist |
|---|---|---|---|---|---|---|---|---|
| Admin | 3% (1-8) | | | | | | | |
| Allied health professional | 1.45 (0.16-13.09) | 2% (1-4) | | | | | | |
| Doctor: consultant | 1.46 (0.15-13.8) | 1.00 (0.15-6.83) | 2% (1-5) | | | | | |
| Doctor: trainee | 2.39 (0.22-26.59) | 1.65 (0.20-13.39) | 1.64 (0.19-14.24) | 1% (0-3) | | | | |
| Healthcare assistant | 0.28 (0.04-1.83) | **0.20* (0.05-0.83)** | **0.20* (0.04-0.90)** | **0.12** (0.02-0.69)** | 9% (6-13) | | | |
| Nurse | 0.51 (0.09-3.09) | 0.35 (0.09-1.37) | 0.35 (0.08-1.49) | 0.21 (0.04-1.15) | 1.80 (0.89-3.65) | 5% (4-7) | | |
| Other: clinical | 1.27 (0.13-12.18) | 0.87 (0.13-6.04) | 0.87 (0.12-6.40) | 0.53 (0.06-4.65) | 4.46 (0.95-20.93) | 2.47 (0.57-10.67) | 2% (1-5) | |
| Physiotherapist | 2.52 (0.06-105.29) | 1.73 (0.05-59.42) | 1.73 (0.05-61.59) | 1.05 (0.03-41.27) | 8.84 (0.31-251.31) | 4.9 (0.18-133.41) | 1.98 (0.06-71.33) | 1% (0-8) |

*p < 0.05; **p < 0.01; ***p < 0.001

Table S5: Post-hoc comparisons for the risk of testing positive for COVID-19 between healthcare worker roles during the second wave. The probability of testing positive for COVID-19 is reported for each healthcare worker role in the diagonal of the matrix (grey cells) with 95% confidence intervals in brackets. The contrasts between roles, as determined by estimated marginal means from a mixed effects logistic regression, are reported as odds ratios with 95% confidence intervals in brackets. Contrasts with statistical significance are in bold.

| Second wave (*Reference level*) | Admin | Allied health professional | Doctor: consultant | Doctor: trainee | Healthcare assistant | Nurse | Other: clinical | Physiotherapist |
|---|---|---|---|---|---|---|---|---|
| Admin | 2% (1-4) | | | | | | | |
| Allied health professional | 1.34 (0.25-7.11) | 1% (1-2) | | | | | | |
| Doctor: consultant | 6.47 (0.66-63.09) | 4.83 (0.59-39.4) | 0% (0-1) | | | | | |
| Doctor: trainee | 1.50 (0.32-6.97) | 1.12 (0.32-3.88) | 0.23 (0.03-1.71) | 1% (1-2) | | | | |
| Healthcare assistant | 0.83 (0.19-3.66) | 0.62 (0.19-2.04) | **0.13* (0.02-0.91)** | 0.56 (0.21-1.50) | 2% (1-3) | | | |
| Nurse | 1.06 (0.27-4.19) | 0.79 (0.28-2.26) | 0.16 (0.03-1.07) | 0.71 (0.31-1.6) | 1.27 (0.62-2.60) | 2% (1-2) | | |
| Other: clinical | 1.67 (0.28-10.05) | 1.25 (0.26-5.95) | 0.26 (0.03-2.34) | 1.11 (0.27-4.61) | 2 (0.51-7.79) | 1.58 (0.45-5.47) | 1% (0-2) | |
| Physiotherapist | 3.95 (0.27-58.14) | 2.95 (0.23-36.99) | 0.61 (0.03-11.94) | 2.63 (0.23-30.32) | 4.73 (0.42-53.13) | 3.73 (0.35-39.17) | 2.37 (0.17-32.59) | 0% (0-2) |

*p < 0.05; **p < 0.01; ***p < 0.001

Table S6. Predictors for the risk of healthcare workers testing positive for COVID-19 during the pandemic. The odds ratio, 95% confidence intervals and statistical significance is reported from models using metrics derived from either 14 days, 7 days or 2 days of data prior to a COVID-19 test being taken. Specific post-hoc comparisons of interest are reported. Results for the models using 7 and 2 days of data are presented in bold if the result is different to that in the models using 14 days of data. For the model using 2 days of data, tests for the physiotherapist staff group were excluded as the records were too few for the model to be stable, and the model for activity on floor 5 was not included as again the model was not stable.

| Predictors | Contrast / Reference | 14 Days (n = 28,909) | 7 Days (n = 26,243) | 2 Days (n = 18,688) |
|---|---|---|---|---|
| Age | - | 0.99 (0.98-1.00) | 0.99 (0.98-1.00) | 0.99 (0.98-1.00) |
| Ethnicity | BAME / White | 1.75 (1.40-2.20)*** | 1.79 (1.42-2.26)*** | 1.66 (1.28-2.15)*** |
| Period | First wave / Summer lull | 2.86 (2.25-3.65)*** | 2.94 (2.29-3.77)*** | 3.31 (2.48-4.42)*** |
| | First wave / Second wave | 9.35 (7.41-11.81)*** | 9.68 (7.59-12.34)*** | 10.92 (8.24-14.46)*** |
| | Summer lull / Second wave | 3.27 (2.53-4.23)*** | 3.29 (2.52-4.30)*** | 3.30 (2.41-4.52)*** |
| Admin | First wave / Summer lull | 5.83 (1.24-27.51)* | 5.04 (1.04-24.51)* | **5.44 (0.66-45.01)** |
| | First wave / Second wave | 9.51 (2.58-35.11)*** | 8.16 (2.14-31.16)*** | 5.13 (1.15-22.78)* |
| | Summer lull / Second wave | 1.63 (0.33-8) | 1.62 (0.33-7.98) | 0.94 (0.13-7.03) |
| Allied health professional | First wave / Summer lull | 7.84 (2.66-23.1)*** | 9.16 (2.92-28.76)*** | 7.85 (2.26-27.26)*** |
| | First wave / Second wave | 11.78 (4.84-28.64)*** | 11.29 (4.61-27.66)*** | 12.5 (4.56-34.27)*** |
| | Summer lull / Second wave | 1.5 (0.45-5) | 1.23 (0.35-4.3) | 1.59 (0.4-6.28) |
| Doctor: consultant | First wave / Summer lull | 4.47 (1.3-15.38)* | 4.5 (1.29-15.64)* | 8.78 (1.8-42.94)** |
| | First wave / Second wave | 32.37 (6.91-151.71)*** | 31.12 (6.59-147.07)*** | 42.22 (6.68-266.73)*** |
| | Summer lull / Second wave | 7.23 (1.28-41.03)* | 6.92 (1.22-39.28)* | **4.81 (0.52-44.06)** |
| Doctor: trainee | First wave / Summer lull | 15.1 (3.92-58.14)*** | 10.91 (3.13-38.03)*** | 8.4 (2.36-29.91)*** |
| | First wave / Second wave | 15.45 (7.16-33.34)*** | 20.56 (8.61-49.09)*** | 32.86 (10.91-98.96)*** |
| | Summer lull / Second wave | 1.02 (0.26-3.98) | 1.88 (0.51-6.96) | 3.91 (0.92-16.69) |
| Healthcare assistant | First wave / Summer lull | 1.51 (0.78-2.92) | 1.73 (0.87-3.44) | **2.27 (1.07-4.82)*** |
| | First wave / Second wave | 7.23 (3.63-14.42)*** | 8.71 (4.21-18.01)*** | 13.13 (5.76-29.95)*** |
| | Summer lull / Second wave | 4.78 (2.53-9.03)*** | 5.03 (2.56-9.85)*** | 5.78 (2.64-12.64)*** |
| Nurse | First wave / Summer lull | 2.25 (1.63-3.11)*** | 2.28 (1.63-3.18)*** | 2.69 (1.83-3.96)*** |
| | First wave / Second wave | 7.57 (5.5-10.42)*** | 7.5 (5.4-10.42)*** | 9.07 (6.21-13.24)*** |
| | Summer lull / Second wave | 3.36 (2.39-4.74)*** | 3.29 (2.32-4.68)*** | 3.37 (2.23-5.09)*** |
| Other: clinical | First wave / Summer lull | 5.19 (1.53-17.57)** | 5.52 (1.63-18.68)** | 6.85 (1.35-34.86)* |

| | | | | |
|---|---|---|---|---|
| | First wave / Second wave | 11.13 (3.78-32.78)*** | 12.63 (4.07-39.17)*** | 6.89 (2.14-22.18)*** |
| | Summer lull / Second wave | 2.15 (0.55-8.37) | 2.29 (0.57-9.25) | 1 (0.19-5.37) |
| Physiotherapist | First wave / Summer lull | 24.65 (1.69-358.89)* | 18.3 (1.24-270.97)* | - |
| | First wave / Second wave | 63.05 (8.31-478.51)*** | 45.15 (5.81-350.57)*** | - |
| | Summer lull / Second wave | 2.56 (0.13-52.19) | 2.47 (0.12-50.14) | - |
| No. of shifts | - / First wave | 1.08 (1.04-1.13)*** | **1.08 (1.00-1.18)** | **0.84 (0.62-1.14)** |
| | - / Summer lull | 1.10 (1.04-1.16)*** | 1.12 (1.01-1.23)* | **1.03 (0.70-1.52)** |
| | - / Second wave | 1.01 (0.96-1.06) | 0.98 (0.90-1.08) | 0.79 (0.54-1.15) |
| No. of night shifts (relative) | - / First wave | 1.05 (0.96-1.15) | **1.14 (1.00-1.29)*** | 0.92 (0.68-1.24) |
| | - / Summer lull | 1.13 (1.03-1.25)** | 1.16 (1.00-1.33)* | **1.02 (0.72-1.44)** |
| | - / Second wave | 1.07 (0.98-1.18) | 1.05 (0.91-1.21) | 0.93 (0.64-1.33) |
| No. of patients (log 2) | - / First wave | 1.29 (1.19-1.41)*** | 1.33 (1.20-1.47)*** | 1.50 (1.30-1.73)*** |
| | - / Summer lull | 1.34 (1.20-1.48)*** | 1.43 (1.27-1.61)*** | 1.51 (1.29-1.77)*** |
| | - / Second wave | 1.18 (1.07-1.30)** | 1.19 (1.07-1.33)*** | 1.22 (1.05-1.42)** |
| No. of COVID-19 patients (log 2; relative) | - / First wave | 0.80 (0.67-0.95)* | 0.77 (0.62-0.97)* | **0.93 (0.67-1.30)** |
| | - / Summer lull | 1.13 (0.82-1.55) | 1.20 (0.82-1.76) | 1.09 (0.56-2.13) |
| | - / Second wave | 1.20 (1.05-1.37)** | 1.27 (1.09-1.49)** | 1.32 (1.06-1.64)* |
| No. of patient contacts (log 2; relative) | - / First wave | 0.83 (0.75-0.91)*** | 0.86 (0.77-0.95)** | 0.87 (0.77-0.98)* |
| | - / Summer lull | 0.92 (0.81-1.04) | 0.97 (0.86-1.10) | 1.10 (0.96-1.27) |
| | - / Second wave | 0.97 (0.86-1.08) | 1.05 (0.94-1.18) | 1.04 (0.91-1.19) |
| No. of COVID-19 contacts (log 2) | - / First wave | 0.77 (0.67-0.89)*** | 0.75 (0.64-0.89)** | 0.72 (0.58-0.90)** |
| | - / Summer lull | 0.90 (0.74-1.09) | 1.01 (0.81-1.26) | 0.87 (0.57-1.32) |
| | - / Second wave | 1.00 (0.88-1.14) | 1.02 (0.89-1.17) | 1.01 (0.85-1.20) |
| No. of floors | - / First wave | 1.06 (0.99-1.13) | 1.06 (0.97-1.15) | 1.11 (0.97-1.28) |
| | - / Summer lull | 0.99 (0.90-1.10) | 1.00 (0.88-1.14) | 0.91 (0.72-1.15) |
| | - / Second wave | 1.04 (0.96-1.13) | 1.08 (0.98-1.19) | 1.06 (0.90-1.25) |
| No. of COVID-19 floors (relative) | - / First wave | 1.52 (1.28-1.81)*** | 1.59 (1.31-1.92)*** | **1.19 (0.93-1.50)** |
| | - / Summer lull | 1.74 (1.40-2.15)*** | 1.75 (1.39-2.21)*** | 1.94 (1.41-2.65)*** |
| | - / Second wave | 1.04 (0.85-1.27) | 0.97 (0.77-1.21) | 0.93 (0.69-1.26) |
| Activity on floor 0 | First wave: Yes / No | 2.04 (1.44-2.89)*** | 2.45 (1.68-3.56)*** | 3.44 (2.19-5.40)*** |

| | | | | |
|---|---|---|---|---|
| | Summer lull: Yes / No | 1.64 (1.04-2.58)* | **1.40 (0.84-2.32)** | **0.95 (0.48-1.91)** |
| | Second wave: Yes / No | 1.26 (0.82-1.92) | 1.19 (0.74-1.92) | 0.96 (0.50-1.86) |
| Activity on floor 1 | First wave: Yes / No | 2.11 (1.42-3.12)*** | 2.33 (1.51-3.58)*** | 2.68 (1.52-4.74)*** |
| | Summer lull: Yes / No | 2.52 (1.57-4.05)*** | 2.81 (1.69-4.67)*** | 4.35 (2.33-8.11)*** |
| | Second wave: Yes / No | 1.35 (0.86-2.13) | 1.23 (0.73-2.06) | 1.31 (0.66-2.62) |
| Activity on floor 2 | First wave: Yes / No | 0.68 (0.43-1.05) | **0.58 (0.36-0.96)*** | 0.69 (0.39-1.24) |
| | Summer lull: Yes / No | 1.33 (0.79-2.25) | 1.14 (0.62-2.08) | 0.58 (0.23-1.48) |
| | Second wave: Yes / No | 1.30 (0.83-2.04) | 1.13 (0.68-1.87) | 0.90 (0.47-1.75) |
| Activity on floor 3 | First wave: Yes / No | 0.39 (0.26-0.58)*** | 0.35 (0.22-0.54)*** | 0.28 (0.16-0.49)*** |
| | Summer lull: Yes / No | 0.45 (0.25-0.79)** | 0.44 (0.24-0.80)** | 0.40 (0.19-0.86)* |
| | Second wave: Yes / No | 0.91 (0.59-1.39) | 0.96 (0.61-1.52) | 1.05 (0.59-1.87) |
| Activity on floor 5 | First wave: Yes / No | 0.82 (0.31-2.16) | 0.91 (0.29-2.84) | - |
| | Summer lull: Yes / No | 1.12 (0.37-3.41) | 1.14 (0.30-4.33) | - |
| | Second wave: Yes / No | 0.60 (0.14-2.51) | 0.27 (0.02-3.03) | - |
| Activity on floor 6 | First wave: Yes / No | 0.90 (0.53-1.54) | 0.55 (0.26-1.14) | 1.10 (0.44-2.75) |
| | Summer lull: Yes / No | 1.26 (0.63-2.50) | 1.53 (0.72-3.26) | **2.91 (1.16-7.28)*** |
| | Second wave: Yes / No | 1.30 (0.80-2.12) | 1.66 (1.00-2.77) | **2.10 (1.11-3.95)*** |
| Activity on floor 7 | First wave: Yes / No | 1.96 (1.31-2.94)*** | 2.64 (1.71-4.09)*** | 3.35 (1.95-5.73)*** |
| | Summer lull: Yes / No | 1.60 (0.94-2.72) | 1.66 (0.93-2.96) | 1.89 (0.90-3.97) |
| | Second wave: Yes / No | 1.20 (0.77-1.86) | 1.07 (0.64-1.77) | 1.19 (0.60-2.36) |
| Activity on floor 8 | First wave: Yes / No | 1.21 (0.80-1.82) | 1.01 (0.63-1.63) | 0.80 (0.40-1.59) |
| | Summer lull: Yes / No | 1.27 (0.75-2.17) | 1.53 (0.86-2.72) | 1.32 (0.55-3.12) |
| | Second wave: Yes / No | 1.32 (0.79-2.19) | 1.33 (0.75-2.36) | 1.00 (0.42-2.37) |
| Activity on floor 9 | First wave: Yes / No | 1.92 (1.23-3.00)** | 1.76 (1.05-2.94)* | **1.66 (0.80-3.46)** |
| | Summer lull: Yes / No | 1.50 (0.84-2.68) | **1.89 (1.01-3.52)*** | **2.51 (1.06-5.98)*** |
| | Second wave: Yes / No | 1.14 (0.69-1.88) | 1.20 (0.69-2.07) | 0.69 (0.30-1.60) |
| Activity on floor 10 | First wave: Yes / No | 2.39 (1.62-3.53)*** | 2.70 (1.75-4.15)*** | 2.10 (1.15-3.85)* |
| | Summer lull: Yes / No | 2.07 (1.28-3.35)** | **1.54 (0.91-2.61)** | **1.00 (0.51-1.96)** |
| | Second wave: Yes / No | 1.13 (0.67-1.93) | 1.33 (0.76-2.34) | 1.14 (0.54-2.38) |
| Activity on floor 11 | First wave: Yes / No | 0.99 (0.55-1.75) | 0.76 (0.35-1.63) | 0.72 (0.26-2.03) |

| | | | | |
|---|---|---|---|---|
| | Summer lull: Yes / No | 0.36 (0.14-0.92)* | **0.35 (0.12-1.01)** | **0.36 (0.08-1.55)** |
| | Second wave: Yes / No | 0.69 (0.35-1.36) | 0.77 (0.37-1.60) | 0.73 (0.28-1.92) |
| Activity on floor 12 | First wave: Yes / No | 0.72 (0.37-1.41) | 0.66 (0.31-1.40) | 0.55 (0.20-1.53) |
| | Summer lull: Yes / No | 0.52 (0.23-1.20) | 0.60 (0.25-1.43) | 0.49 (0.15-1.57) |
| | Second wave: Yes / No | 0.82 (0.45-1.47) | 0.99 (0.54-1.81) | 0.90 (0.41-1.97) |
| Activity on floor 13 | First wave: Yes / No | 0.76 (0.50-1.14) | 0.66 (0.42-1.04) | 0.73 (0.42-1.29) |
| | Summer lull: Yes / No | 0.45 (0.23-0.89)* | **0.55 (0.27-1.12)** | **0.79 (0.35-1.80)** |
| | Second wave: Yes / No | 0.74 (0.42-1.30) | 0.74 (0.39-1.41) | 0.65 (0.26-1.59) |
| Activity on floor 14 | First wave: Yes / No | 0.80 (0.53-1.20) | 0.79 (0.51-1.24) | 0.87 (0.50-1.51) |
| | Summer lull: Yes / No | 0.33 (0.15-0.73)** | 0.38 (0.16-0.92)* | **0.27 (0.06-1.14)** |
| | Second wave: Yes / No | 0.73 (0.41-1.29) | 0.75 (0.39-1.43) | 0.60 (0.24-1.48) |
| Activity on floor 15 | First wave: Yes / No | 0.26 (0.04-1.62) | 0.39 (0.06-2.53) | 0.57 (0.04-7.82) |
| | Summer lull: Yes / No | 0.66 (0.11-4.08) | 0.97 (0.15-6.19) | 2.27 (0.34-15.11) |
| | Second wave: Yes / No | 1.88 (0.96-3.68) | **2.38 (1.16-4.88)*** | **3.09 (1.22-7.79)*** |
| Activity on floor 16 | First wave: Yes / No | 0.66 (0.43-1.00) | **0.54 (0.34-0.88)**** | 0.56 (0.30-1.02) |
| | Summer lull: Yes / No | 0.37 (0.19-0.72)** | 0.29 (0.13-0.65)** | 0.31 (0.11-0.88)* |
| | Second wave: Yes / No | 0.61 (0.33-1.14) | 0.63 (0.32-1.27) | 0.65 (0.26-1.60) |
| No. of door events (log 2) | - / First wave | 1.04 (0.97-1.12) | 1.05 (0.97-1.13) | **1.13 (1.03-1.24)*** |
| | - / Summer lull | 1.03 (0.95-1.11) | 1.02 (0.94-1.10) | **1.15 (1.03-1.29)*** |
| | - / Second wave | 1.03 (0.96-1.11) | 1.04 (0.96-1.12) | 1.02 (0.92-1.13) |
| No. of COVID-19 door events (log 2; relative) | - / First wave | 1.12 (1.05-1.19)*** | 1.08 (1.00-1.15)* | **1.04 (0.94-1.14)** |
| | - / Summer lull | 1.15 (1.08-1.23)*** | 1.15 (1.07-1.24)*** | 1.19 (1.08-1.32)*** |
| | - / Second wave | 1.02 (0.95-1.10) | 1.03 (0.95-1.12) | 0.99 (0.89-1.11) |

*p < 0.05; **p < 0.01; ***p < 0.001
